# Performance assessment of large language models in cancer staging: Comparative analysis of Mistral models

**DOI:** 10.1101/2025.09.10.25335246

**Authors:** Roman Rouzier, Valentin Harter, Ethan Rouzier, Victor Ferment, Simon Gruau, Benoit Andre, Cécile Saumon-Sud, Lawrence Nadin, Aurélien Corroyer-Dulmont, Nicolas Vigneron

**Author notes:** Corresponding author: Pr Roman ROUZIER, Comprehensive Cancer Center François Baclesse, 3 avenue du Général Harris, Caen, 14000, France.

## Abstract

Cancer staging plays a critical role in treatment planning and prognosis but is often embedded in unstructured clinical narratives. To automate the extraction and structuring of staging data, large language models (LLMs) have emerged as a promising approach. However, their performance in real-world oncology settings has yet to be systematically evaluated. Herein, we analysed 1000 oncological summaries from patients receiving treatment for breast cancer between 2019 and 2020 at the François Baclesse Comprehensive Cancer Centre, France. Five Mistral artificial intelligence–based LLMs were evaluated (i.e. Small, Medium, Large, Magistral and Mistral:latest) for their ability to derive the cancer stage and identify staging elements. Larger models outperformed their smaller counterparts in staging accuracy and reproducibility (kappa > 0.95 for Mistral Large and Medium). Mistral Large achieved the highest accuracy in deriving the cancer stage (93.0%), surpassing the original clinical documentation in several cases. The LLMs consistently performed better in deriving the cancer stage when working through tumour size, nodal status and metastatic components compared to when they were directly requested stage data. The top-performing models had a test–retest reliability exceeding 97%, while smaller models and locally deployed versions lacked sufficient robustness, particularly in handling unit conversions and complex staging rules. The structured, stepwise use of LLMs that emulates clinician reasoning offers a more efficient, transparent and reproducible approach to cancer staging, and the study findings support LLM integration into digital oncology workflows.

## 1. Introduction

Cancer staging is a critical component in the diagnosis and treatment planning of oncological diseases. Accurate staging helps clinicians establish an accurate prognosis and select the most appropriate therapeutic strategies. However, the extraction and structuring of staging information from unstructured medical records remain a persistent challenge in oncology. Staging is primarily based on the TNM classification, which is based on tumour size and extent (T), regional lymph node involvement (N), and the presence or absence of distant metastases (M). The necessity of automated structured data extraction from clinical records echoes earlier works involving the application of deep learning to electronic health records (Rajkomar et al., 2018). Clinicians often document patient information in narrative form, which can create inconsistencies and difficulties in data retrieval and analysis (Bilal et al., 2025).

Beyond its clinical role, cancer staging has also been considered a cornerstone of quality assurance in oncology. For example, the Organisation of European Cancer Institutes (OECI) has developed a comprehensive accreditation programme that relies on the systematic collection and analysis of quality and safety of care indicators across comprehensive cancer centres. These indicators, which include process, vigilance and outcome measures, are essential for monitoring the pertinence and safety of care pathways. In this context, accurate stratification of patients according to organ involvement and tumour stage is critical for not only guiding therapeutic decisions but also ensuring comparability of outcomes across institutions. Reliable staging information facilitates survival assessment according to stage, benchmarking of waiting times and the evaluation of adherence to evidence-based guidelines. Consequently, automating the extraction and structuring of such staging data from clinical narratives facilitates accreditation processes and the continuous improvement of cancer care delivery in Europe, which is directly aligned with the OECI quality agenda. This need to reliably retrieve staging information for quality assessment and accreditation purposes forms the basis of our study.

Recently, the advent of artificial intelligence (AI), particularly large language models (LLMs), has opened new avenues for addressing these challenges. LLMs, which are capable of understanding and generating human-like text, offer promising solutions for transforming unstructured medical data into structured formats (Vrdoljak et al., 2025). This transformation is crucial for enhancing data analysis efficiency, improving patient care and facilitating research. More broadly, LLMs have already demonstrated their potential in the automatic summarisation of biomedical text by supporting literature screening, medical knowledge management and the clinical narrative structuring (Huang et al., 2025). This growing body of work underscores the importance of systematically evaluating their application in the field of oncology.

Therefore, this study aimed to evaluate the performance of advanced LLMs in extracting and structuring cancer staging data from unstructured medical texts. These models are available in various sizes corresponding to the number of parameters, which could potentially influence their performance and computational requirements (Tinn et al., 2023; Touvron et al., 2023). To date, however, systematic analyses comparing the performance of models according to their size and usage strategy, particularly in clinical applications such as cancer staging, have been limited. This evaluation is essential for understanding the potential application of AI in clinical settings and improving cancer data management.

This study contributes to the growing research on AI in healthcare, particularly in the field of oncology. By comparing the performance of Mistral AI’s LLMs, we aim to highlight their strengths, limitations and potential.

## 2. Related Work

The use of LLMs in clinical natural language processing (NLP) has grown exponentially over the past few years. According to Yu et al. (2024), over 1,698 peer-reviewed studies on LLMs in biomedical and health informatics were published between January 2022 and December 2023, reflecting a sharp rise from fewer than 100 before 2020. Tripathi et al. (2025) demonstrated that locally deployed LLMs with consensus-based reasoning achieved high agreement in extracting structured diagnostic data, including cancer staging, from pathology reports. Their approach highlighted the benefits of separating extraction from interpretation to improve reliability and interpretability of results.

Using untrained models can emphasise their ability to deliver satisfactory results while offering a more universal approach. This method is consistent with the prevailing trend in clinical NLP wherein a common reliance on general-purpose LLMs have been noted without fine-tuning. For instance, Chen et al. (2025), who reviewed 24 oncology-focused NLP studies, reported that 67% used fine-tuning strategies, whereas 21% relied on prompt engineering. However, Shool et al. (2025) found that only 6.45% of the models evaluated in over 700 clinical studies were pretrained or fine-tuned to process medical-domain data, whereas 93.55% relied on off-the-shelf LLMs like GPT-4 [10]. Nonetheless, recent evidence from Tozuka et al. (2025) demonstrated that retrieval-augmented generation models, such as NotebookLM, could outperform GPT-4 in lung cancer staging tasks. This finding reinforces the notion that integrating structured domain knowledge, whether through retrieval systems or fine-tuning, can significantly enhance LLM performance on clinical reasoning tasks.

In addition to the training strategy, the model size consistently showed a strong influence on performance. Lee et al. (2025) noted that smaller models face limitations when handling granular pathology details, particularly measurements and staging data [12]. However, this observation requires further confirmation and precise quantification. Several studies have emphasised the importance of reasoning emulation in clinical NLP. Levine et al. (2024) showed that GPT-3 can self-assess its diagnostic confidence, which facilitates more reliable human–AI collaboration. Moreover, recent multilingual evaluations of GPT-4 and other LLMs suggest their robustness across languages, an essential feature for their deployment in non-English clinical environments (Menezes et al., 2025). This linguistic universality is particularly promising for cross-border healthcare applications; however, it may inherently favour larger-scale models, an aspect that warrants further investigation. Although prior work has demonstrated the general feasibility of LLMs in extracting tumour size, nodal status and metastatic (TNM) or American Joint Committee on Cancer stages from pathology reports, only a few studies have systematically benchmarked models of varying sizes or explored how structured versus direct prompting affects staging accuracy and reproducibility in a broad range of clinical documents. Saluja et al., 2025 investigated the use of large language models (LLMs) to automatically extract key information on cancer type and stage from pathology reports. Their study demonstrates the technical feasibility and high performance of these models when applied to a relatively homogeneous corpus of data. However, it is important to note that the results rely exclusively on pathology reports. While rich in clinical content, these documents are produced according to relatively standardized formats, with more structured language and controlled vocabulary compared to other medical sources. This homogeneity facilitates the task for LLMs and likely contributes to the strong results observed. By contrast, extrapolating these findings to more heterogeneous data—such as clinical notes, radiology reports, or free-text documentation—is less straightforward. These sources display far greater linguistic and contextual variability, which makes automated information extraction by AI significantly more challenging.

Our study directly addresses this gap by comparing Mistral models across deployment modes, parameter sizes and prompting strategies for providing novel insights into performance trade-offs and clinical utility. This benchmarking is especially relevant in the current context of environmental sustainability, given the significant variation in energy consumption and carbon footprint across LLM architectures and deployment strategies. As emphasised by Gaetani et al. (2024), the growing adoption of AI in medicine must be balanced with responsible and energy-efficient practices, particularly when deploying large-scale models for routine clinical use.

In summary, although the literature suggests that LLMs could be used for extracting staging data from pathology reports, more studies that specifically compare model sizes and apply these techniques to a broader range of clinical documents are needed. This approach provides a more comprehensive understanding of the capabilities and limitations of LLMs in clinical NLP.

## 3. Research Objectives

This study aimed to systematically evaluate the performance of LLMs in extracting and structuring cancer staging data from unstructured medical texts. Specifically, we aimed to assess how variations in model size, ranging from 7 to 123 billion parameters, influenced the accuracy, reliability and efficiency of staging information extraction, including TNM classification, tumour size and metastatic status. By comparing locally deployed models with those accessed via dedicated APIs, we sought to understand the impact of different deployment strategies on performance in clinical settings.

Our central objective was to investigate the role of prompting strategies in determining the quality and consistency of LLM outputs. Accordingly, we compared direct extraction approaches with structured prompting techniques and explicit reasoning and subsequently evaluated their effects on concordance with expert-validated staging data. Through test–retest analyses, we further examined the reproducibility of results across different models and prompting methods.

Ultimately, this study aimed to identify the strengths and limitations of LLMs in real-world oncological applications by quantifying the agreement between LLM-generated staging and reference data from expert panels or clinical databases while also analysing systematic errors, such as misinterpretation of TNM criteria or measurement inaccuracies, to identify opportunities for model refinement. Our findings provide actionable insights for clinicians and researchers and support the integration of LLMs into oncological practice by focusing on accuracy and clinical utility.

## 4. Proposed Approach

### 4.1. Data Source

This study utilised an observational dataset collected at the François Baclesse Comprehensive Cancer Centre (Caen, Normandy, France), which included 1,000 consecutive patients treated for breast cancer between 2019 and 2020. Data were extracted from oncological summaries contained in the electronic health records. These summaries have been continuously updated by clinicians at each step of the patient’s journey (e.g. consultations and multidisciplinary tumour boards) and serve as a record of the patient’s clinical history. Given that these summaries are authored by multiple physicians, they appear naturally heterogeneous in content and style.

The summaries are unstructured natural language texts, with an average length of 170 words (median: 147; interquartile range: 108), that systematically describe the clinical presentation, treatment management and disease progression. Summaries usually did not include pre-established TNM categories or stage labels. However, constituent elements of the TNM staging are consistently available for TNM classification and cancer staging, even when not explicitly formalised (e.g. ‘15 mm tumour, axilla free’ instead of ‘T1N0’). Examples of these summaries are provided in Supplementary Table 1.

The corpus comprises 6,034 distinct words, with a high proportion of technical terms, reflected by a mean LIX readability score of 71.2 (median: 59). On average, 22.2% of tokens were stop words and 19.1% were numbers (Kwichmann et al., 2025), highlighting the technical and specialised nature of these texts.

All documents were pseudonymised according to standard protocols by removing personal identifiers (e.g. names, dates and patient identification) and stored as plain text files. Ethical approval was granted by the local ethics committee (Centre Francois Baclesse Institutional Review Board #2025-13) and conducted in accordance with French data protection regulations.

### 4.2. Model Selection

This study employed models from Mistral AI (Large, Medium, Small, Magistral and Mistral:latest [f974a74358d6], hereinafter called Mistral Local) to evaluate their performance in structuring cancer staging data. The number of parameters varied across these models, ranging from 7 billion for Mistral Local, to 22 billion for Mistral Small, and reaching 123 billion for Mistral Large. These models were selected for their advanced capabilities in understanding and generating human-like text, which we believe makes them suitable for extracting structured information from unstructured medical texts. The cancer stage determined by Mistral Large was also tested with a prompt asking to justify staging. A dedicated API to specific servers (Mistral Large, Medium, Magistral and small 3.1) and a local server (Mistral:latest, f974a74358d6) were used. As illustrated in Figure 1, the oncological summaries were utilised to prompt the determination of cancer stages, specifically Stages 0, I, II, III and IV, along with the constituent elements of the stage, including TNM classification, maximal tumour size in millimetres, initial metastatic status and the presence of pure ductal carcinoma *in situ* without infiltration. For each patient, the constituent elements of the TNM staging system were used to algorithmically determine a ‘derived stage’ according to American Joint Commission on Cancer guidelines. These data were then compared to a prospectively manually recorded database (CFB Breast database) and to a consolidated staging established by an expert panel (breast tumour board) in case of discordance between the CFB Breast database and Mistral Large model. The pathologic stage (surgical stage) was considered unless the patient received neoadjuvant chemotherapy, in which case, the clinical stage was considered.

**Figure 1.**
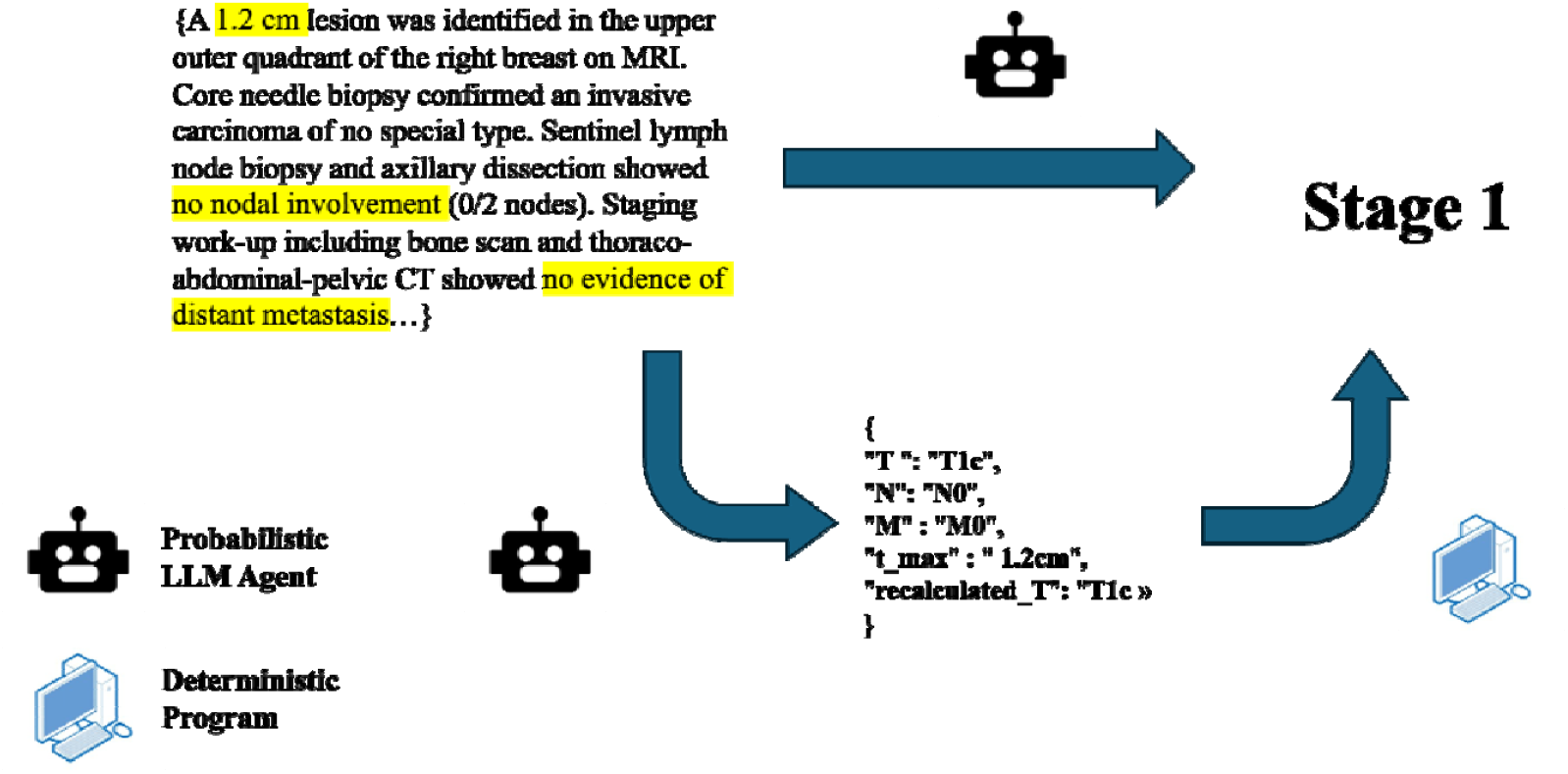
Methodological comparison of LLM-based cancer staging: Direct extraction vs. TNM feature analysis using Mistral AI models

### 4.3. Experimental Setup and Evaluation Metrics

Oncological summaries were input into the models to extract staging information. Each prompt was executed twice to perform a test–retest analysis. The results of the two iterations were compared between the models and correlated with staging data from the CFB Breast database. To determine the stage using Mistral Large with justification, the stage was extracted in the conclusion of the LLM response.

Execution times needed to process cancer stage queries derived from oncological summaries using each model from Mistral AI were recorded. To assess the accuracy and reliability of the models, the staging determined by the prompt and the derived stages were evaluated for concordance with the stage consolidated by the expert. Accuracy, precision, recall and F1 score were evaluated as performance metrics. Results were presented as overall concordance for all stages and as a matrix to evaluate concordance for each stage category. This approach allowed for a comprehensive assessment of the models’ ability to accurately and consistently determine cancer stages from unstructured medical texts.

## 5. Results

The median process execution times were 1.47 s (SD = 1.42 s), 1.46 s (SD = 1.93 s), 0.71 s (SD = 0.34 s) and 1.48 s (SD = 0.91 s) for Mistral Large, Mistral Medium, Mistral Small and Mistral Magistral, respectively. Mistral Large with justification required a significantly longer process execution time (median, 5.80 s; SD = 2.45 s) than did Mistral Large. Similarly, Mistral Local required a significantly longer process execution time (median, 12.1 s; SD = 9.25 s) than did Mistral Large. Interestingly, the maximal process execution times were 12.8, 18.1, 4.84 and 15.7 s for Mistral Large, Mistral Medium, Mistral Small and Mistral Magistral, respectively. The process execution times are summarised in Figure 2. All processing steps were completed quickly, typically within seconds. However, the justification process required more time due to the increased token volume. Although the execution of Mistral Local was comparatively slower, all processing times remained within acceptable operational limits.

**Figure 2.**
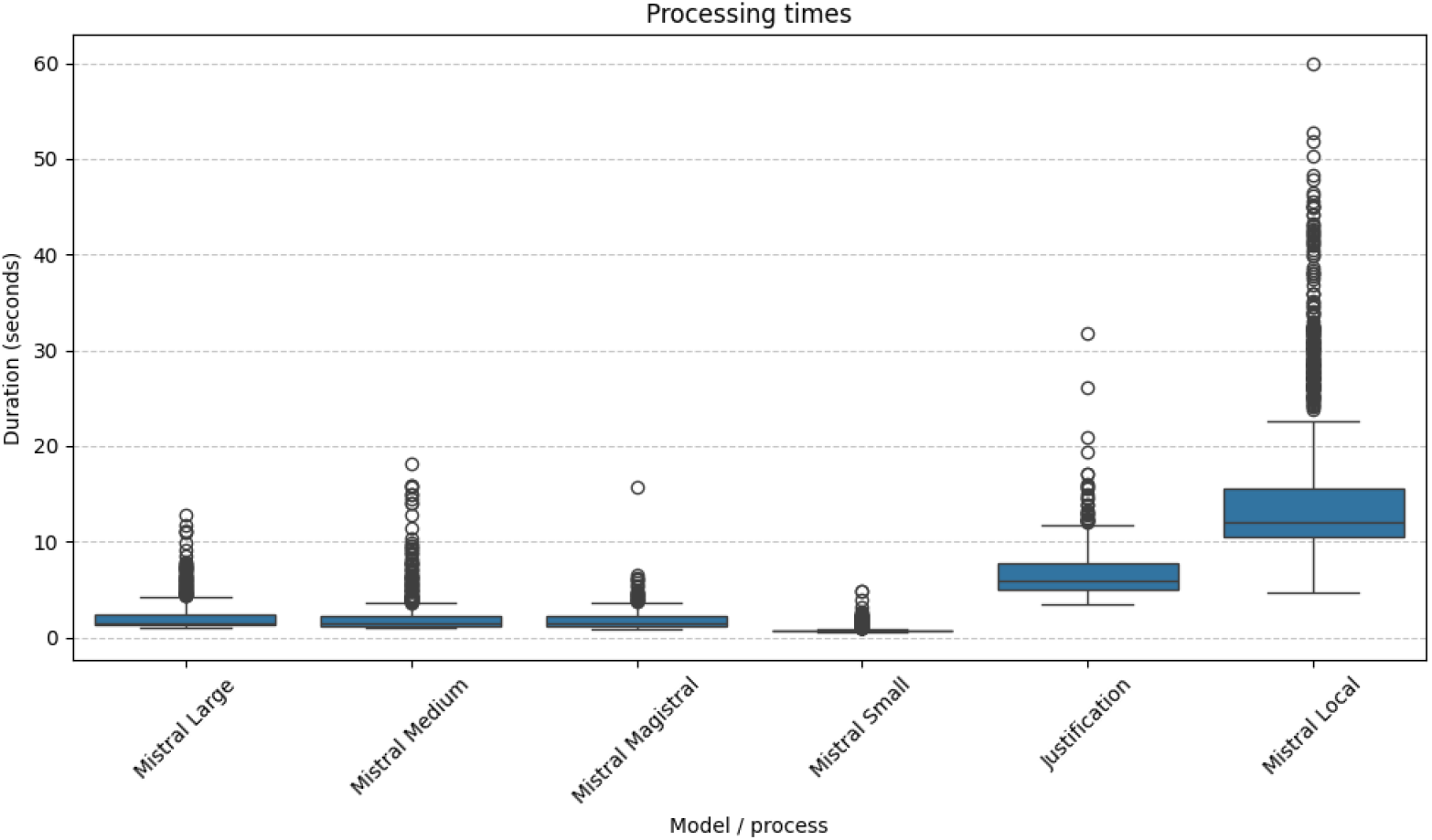
Comparison of execution times including stage and cancer stage query processing from oncological summaries using Mistral models

We evaluated the T, N and M classifications as well as the maximal tumour size and recalculated the T classification to determine potential reasons for misclassification (Figure 3). The smaller models, including the Mistral Local model, struggled to accurately determine the T size or maximal tumour size. These models performed poorly when converting measurements between millimetres and centimetres, unlike the larger models.

**Figure 3.**
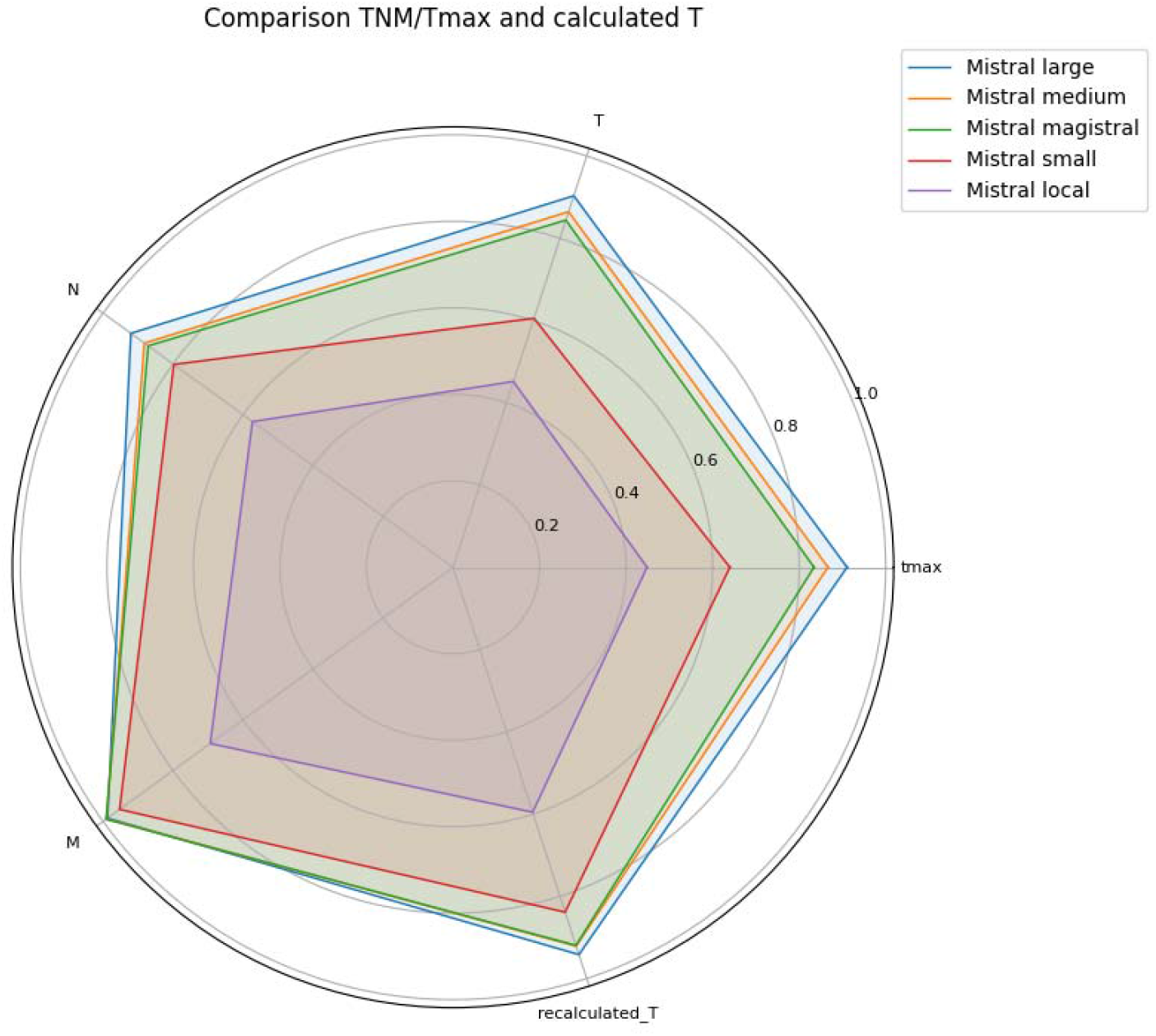
Comparison of TNM classifications, maximal tumour size and recalculated T stages across different Mistral models relative to expert-curated reference data (tumour board) to derive cancer stages.

A test–retest evaluation was then conducted to assess the consistency of staging by the models from Mistral AI (Table 1). For direct stage determination, Mistral Medium and Mistral Large achieved a test–retest reliability rate and kappa of >0.95, whereas Mistral Magistral presented a kappa of <0.95 and Mistral Small presented a reliability rate and kappa of <0.95. For derived stage determination, both Mistral Medium, Mistral Large and Mistral Magistral presented a reliability rate and kappa of >0.95, whereas Mistral Small presented a reliability rate and kappa of <0.95.

**Table 1.**
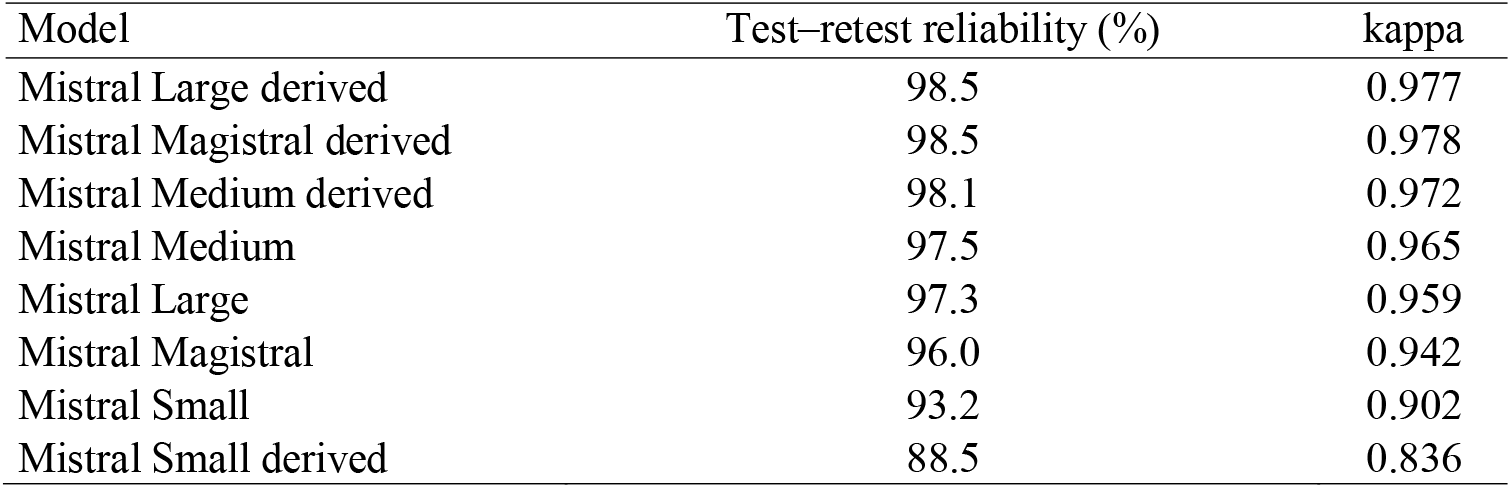
Test–retest reliability rates and kappa coefficients for cancer staging consistency across models from Mistral AI

The test–retest reliability rates and kappa coefficients were relatively high, with the largest models demonstrating clearly superior reproducibility.

The performance of various Mistral models in determining cancer stages from unstructured medical texts was evaluated based on their agreement rates with expert staging. The results are presented in terms of agreement quality, categorised as high (≥90%), moderate (≥70% and <90%) and low (<70%) (Figure 4).

**Figure 4.**
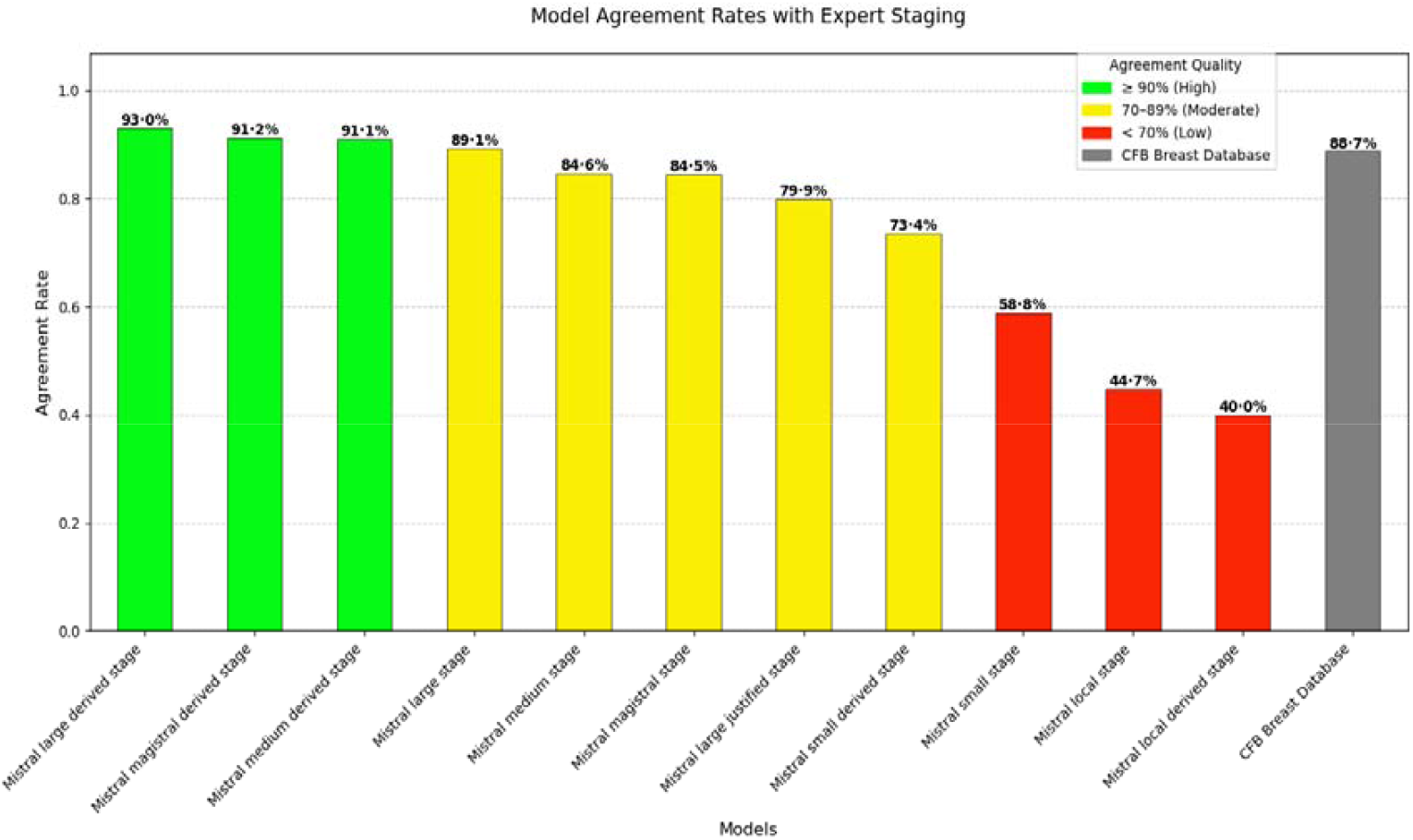
Agreement rates among Mistral models with expert staging for cancer stage determination

Larger Mistral models consistently demonstrated higher performance than did smaller models, with agreement rates increasing proportionally with model size. This finding suggests that larger models were better at capturing complex clinical nuances required for accurate staging. Interestingly, in several cases, the LLM-derived stages were more consistent with clinical records compared to the originally documented stages, highlighting the ability of LLMs to structure unstructured clinical data more effectively. Surprisingly, when prompted to justify its reasoning, Mistral Large exhibited lower accuracy.

Table 2 presents the performance metrics of various Mistral models in determining cancer stages relative to the INS database and expert annotations. The metrics accuracy, precision, recall and F1-score were used to evaluate the effectiveness of each model in capturing the clinical nuances necessary for accurate cancer staging. Notably, larger models, such as Mistral Large, demonstrated superior performance across all metrics, indicating their enhanced capability to interpret complex clinical data. The ‘derived stage’ lines represent algorithmically determined stages, which, in many cases, show improved consistency with clinical records over the originally documented stages.

**Table 2.** Performance metrics of Mistral models in cancer staging prediction

| Model/algorithm | Accuracy | Precision | Recall | f1-score |
| --- | --- | --- | --- | --- |
| Mistral Large derived | 0.93 | 0.932 | 0.93 | 0.929 |
| Mistral Magistral derived | 0.912 | 0.914 | 0.912 | 0.912 |
| Mistral Medium derived | 0.911 | 0.913 | 0.911 | 0.911 |
| Mistral Large | 0.891 | 0.889 | 0.891 | 0.886 |
| Mistral Medium | 0.846 | 0.866 | 0.846 | 0.849 |
| Mistral Magistral | 0.845 | 0.869 | 0.845 | 0.847 |
| Mistral Large with justification | 0.8 | 0.757 | 0.8 | 0.77 |
| Mistral Small derived | 0.734 | 0.772 | 0.734 | 0.743 |
| Mistral Small | 0.588 | 0.679 | 0.588 | 0.588 |
| Mistral Local | 0.523 | 0.683 | 0.523 | 0.529 |
| Mistral Local derived | 0.4 | 0.68 | 0.4 | 0.474 |
| *Record from CFB breast database* | *0.887* | *0.881* | *0.887* | *0.883* |

We then evaluated the stage predictions using a matrix that compares the model outputs to expert-annotated stages to assess potential biases in accuracy. Prediction accuracy was not skewed. Figure 5 presents the results for the Mistral Large model-derived stages. Similar findings were observed across other Mistral model variants (data not shown).

**Figure 5.**
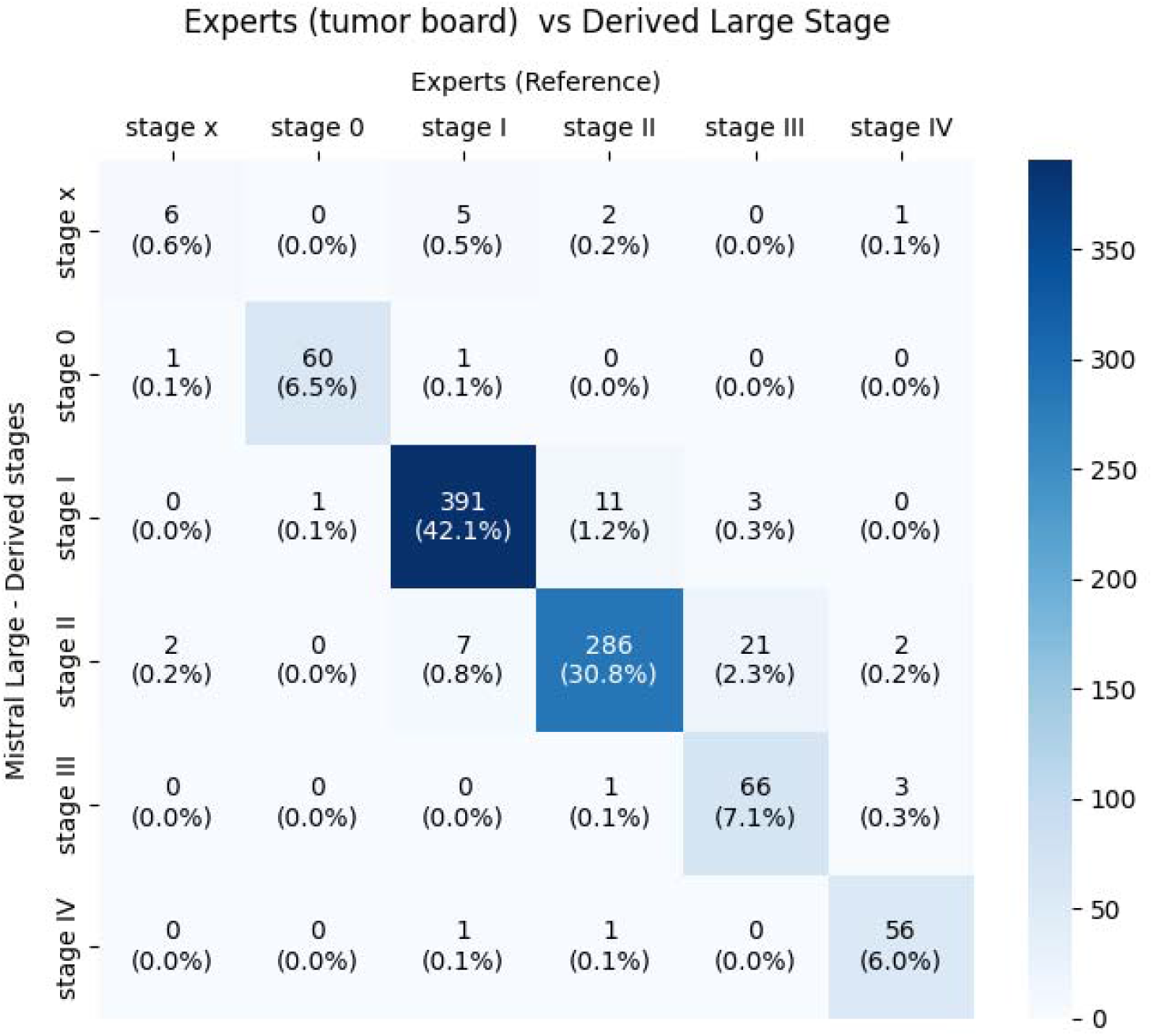
Confusion matrix: Comparison between expert staging and Mistral Large-derived stage predictions

## 6. Discussion

The application of LLMs in healthcare, particularly in oncology, presents a transformative opportunity to enhance the structuring and analysis of unstructured medical data. The current study evaluated the performance of various Mistral AI models in extracting and structuring cancer staging data from unstructured clinical texts, revealing significant insights into their potential and limitations in real-world clinical settings. Our findings suggest that achieving the highest level of accuracy requires the use of a very large-capacity models in combination with structured extraction of clinical information and the application of medical expert-informed algorithms, such as TNM-based staging logic, to ensure consistency, reliability and clinical validity.

Overall, our findings demonstrated that larger models had consistently better accuracy and reliability for staging predictions than did their smaller counterparts. This aligns with previous research showing a positive correlation between model capacity and clinical task performance, likely due to enhanced ability to capture complex semantic and contextual cues within medical narratives (Tinn et al., 2023). The higher concordance rates and kappa coefficients observed in larger Mistral AI models further support their suitability for tasks requiring nuanced interpretation, such as cancer staging. However, the generalizability of our results may come into question given the use of French in our study. Although LLMs are mostly trained on an English corpus, recent studies, and particularly the largest one, have shown the robustness of LLMs across multilingual clinical note processing (Menezes et al., 2024). Our study occasionally observed mixed-language outputs with smaller models, especially in binary yes/no responses that alternated between French and English (data not shown). This finding strongly suggests that larger models exhibit superior multilingual capabilities and that model size may play a critical role in ensuring linguistic consistency and performance in non-English clinical environments.

Our results are consistent with those of Tripathi et al. (2025), who implemented a locally deployed, consensus-based LLM architecture across thousands of pathology reports. In particular, their work confirmed high agreement levels (above 90%) for most diagnostic variables, including cancer stage, using multiple models and expert-validated reasoning mechanisms. This finding underscores the necessity of emulating human reasoning to improve the reliability and interpretability of LLMs, particularly in complex clinical tasks characterised by inherent ambiguity. This approach critically relies on the LLMs’ ability to transform unstructured medical data into coherent and analysable representations, a prerequisite that we had demonstrated in our study, which constitutes its principal added value. Separating the extraction and prediction tasks improved performance by rendering the staging process deterministic, with only the extraction step remaining probabilistic. The present study also revealed challenges, particularly with smaller models that struggled with measurement interpretation (e.g. tumour size conversion between units). This finding had been corroborated by prior observations from Lee et al. (2025), who highlighted decreased model accuracy when handling histologic and staging data from thyroid pathology reports. These issues emphasise the importance of model selection relative to the granularity and precision required by the clinical task.

In the current study, models determined the stage based on an oncological summary that synthesises medical information, whereas the hospital breast cancer database was compiled from the entire patient file, which means it could provide staging information that differs from that in the oncological summary. We observed occasionally inaccuracies in the oncological summaries authored by clinicians, especially when dictated using voice recognition systems. Such discrepancies can often be explained by inconsistencies between clinical and imaging assessments, as well as the variable predictive value and limited reproducibility of clinical examinations alone. Despite these inherent limitations in the reference data, our study remains highly informative. Moreover, it contributes valuable insights by comparing multiple model configurations, implementing a structured evaluation strategy and assessing reliability metrics that appear largely unaffected regardless of whether the stage was derived from the entire medical record or from a synthesised oncological summary. Looking ahead, LLMs that can process and analyse the entirety of a patient’s documents can be leveraged to provide a more comprehensive and cohesive synthesis of medical information, potentially reducing discrepancies and improving the accuracy of staging information. However, supplying several different documents to the models could increase calculation times given the need to produce a synthesis by cross-referencing potentially discordant information. Similarly, this more complex task could challenge these models, potentially affecting their performance levels. As such, LLMs need to have dedicated confidence scores to distinguish difficult cases that could be referred to human experts.

Regarding operational feasibility, although local execution of LLMs involved longer processing times, they remained within acceptable limits. This finding suggests that with appropriate infrastructure, real-time or near-real-time deployment is technically viable. The current study was nonetheless limited by the pseudonymisation process, particularly the modification of dates, which may hinder the identification of de novo metastatic cases— information that is crucial for clinical interpretation. Our findings highlight the importance of developing secure and trusted infrastructures that facilitate the use of data in clinical settings, in line with the requirements of the forthcoming European Artificial Intelligence Act. These infrastructures include the implementation of certified environments, such as cloud services with appropriate security certifications, and the deployment of high-performance computing resources (e.g. servers and GPUs) to support clinically relevant AI applications, whether on-site or remotely. As emphasised in recent work advocating for on-premise LLM use (Tripathi et al., 2025), locally deployed models help mitigate privacy concerns associated with cloud-based services, offering a safer framework for clinical environments subject to regulatory constraints. However, the lower performance of locally deployed models observed in our study highlights the trade-off between privacy and computational efficiency, suggesting the need for further optimisation to enhance their effectiveness in clinical settings.

Our findings have both theoretical and practical implications. We show that LLMs can emulate medical reasoning when constrained by domain knowledge and deterministic logic (e.g., TNM algorithms). Unlike approaches based on retrieval-augmented generation, knowledge graphs, graph neural networks, prompt chaining/chain-of-thought, or fine-tuning (Delleani et al., 2024; Gajo & Barrón-Cedeño, 2025; Bai et al., 2025; Kim et al., 2024), we decouple probabilistic extraction from a rule-based staging engine. This is conceptually close to Saluja et al. (2025) on pathology-based staging; importantly, they themselves call for adding domain-specific heuristics and structured reasoning to reduce AJCC misclassifications. We operationalize that recommendation via a deterministic algorithmic layer that constrains outputs, enables automatic checks against clinical guidelines, and lowers computational cost compared with fully LLM-driven pipelines. The resulting controlled hybrid improves reproducibility, interpretability, and error attribution. While recent NER systems often rely on complex prompting that increases token usage and engineering overhead (Wang et al., 2025), our pipeline uses a single LLM for extraction, with deterministic rules ensuring staging consistency—an efficient and scalable design for healthcare deployment (Tsaneva et al., 2025). From a practical standpoint, our results show that LLMs can be feasibly integrated into oncology workflows and can address concrete institutional needs, such as accreditation (e.g. OECI self-assessment) and quality monitoring (e.g. waiting times). The ability of larger models to consistently outperform smaller ones underscores the importance of appropriate model selection according to the granularity and precision required by the clinical task. Importantly, the heterogeneity of oncological summaries in our dataset mirrors information available in real-world clinical settings, wherein multiple physicians contribute to non-standardised documentation. Our work highlights the potential of LLMs for multicentre harmonisation, benchmarking and federated data infrastructures by demonstrating that they can reliably extract and reconstruct structured information from such heterogeneous inputs.

In conclusion, the current study demonstrates the potential of LLMs in oncology and contributes to the expanding body of literature on this matter. In particular, our findings shown that compared to smaller and locally deployed models, larger language models demonstrated superior accuracy, efficiency and consistency of cancer staging extraction from unstructured clinical texts. However, this improvement comes at the expense of substantially greater computational and energy requirements, raising important concerns regarding sustainability and scalability in clinical practice (Gaetani et al., 2025). Future research should therefore prioritise integration into clinical workflows, examine real-world performance on diverse datasets and address limitations inherent to smaller models. Our study also highlights the need to build on medical expertise and emulate human reasoning to enhance the transparency, safety and clinical relevance of these outstanding tools.

### Data sharing

De-identified data and code may be available for educational or research purposes upon reasonable request to the Principal Investigator Roman Rouzier, subject to interinstitutional data sharing agreements.

### Use of AI

During the preparation of this work the authors used Mistral AI to improve the language and clarity of the text. After using this tool, the authors reviewed and edited the content as needed and take full responsibility for the content of the publication.

---

**Table S1.**
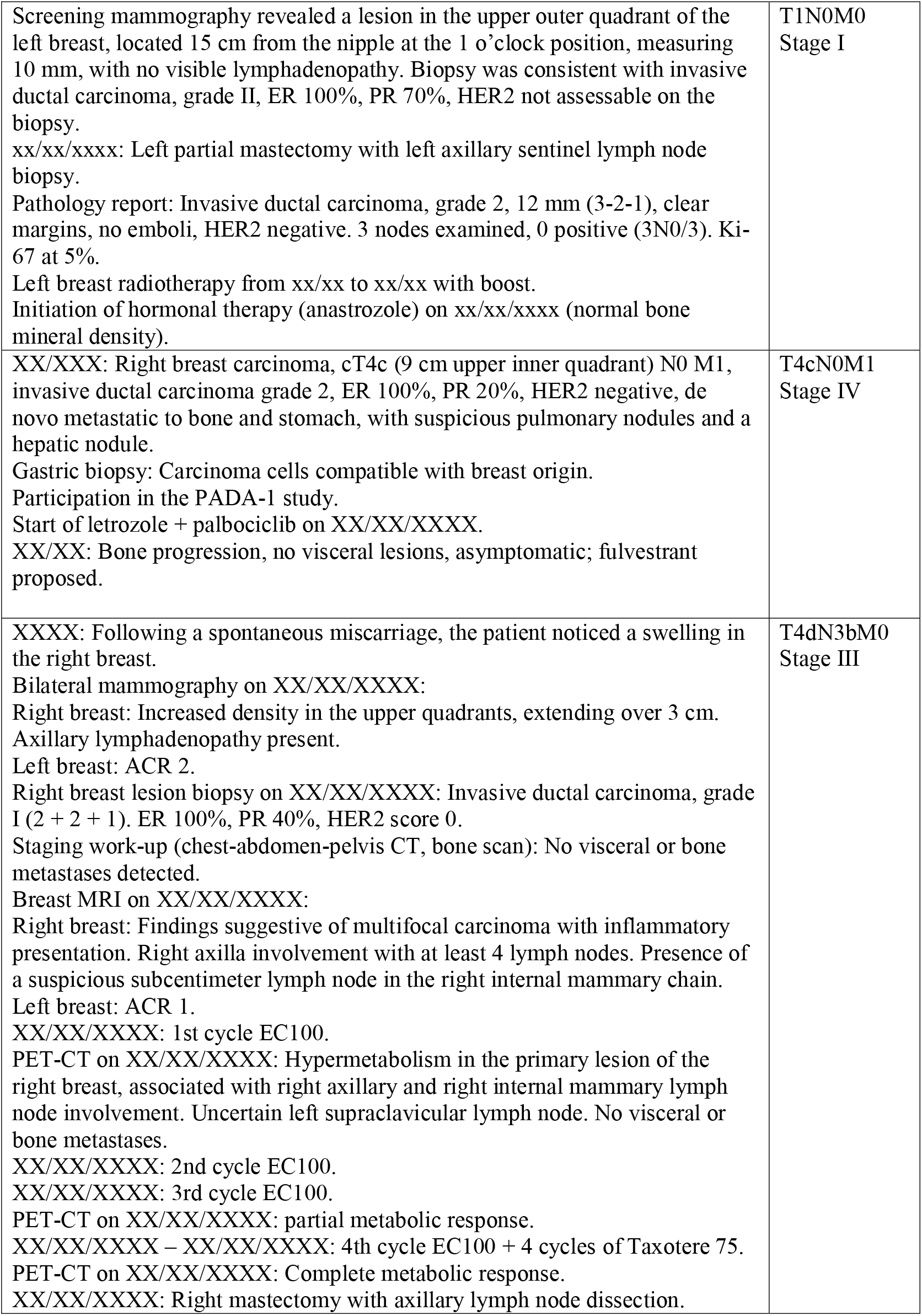
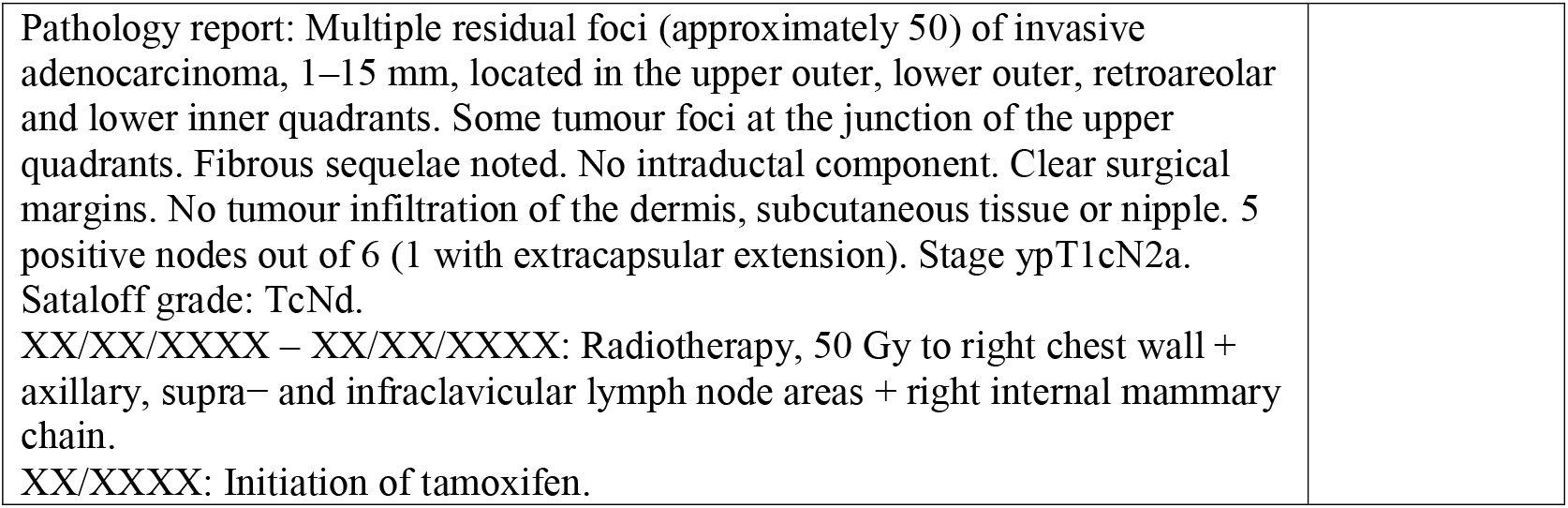
Supplemental Examples of oncological summaries

|  |  |
| --- | --- |
| Screening mammography revealed a lesion in the upper outer quadrant of the left breast, located 15 cm from the nipple at the 1 o'clock position, measuring 10 mm, with no visible lymphadenopathy. Biopsy was consistent with invasive ductal carcinoma, grade II, ER 100%, PR 70%, HER2 not assessable on the biopsy.<br>xx/xx/xxxx: Left partial mastectomy with left axillary sentinel lymph node biopsy.<br>Pathology report: Invasive ductal carcinoma, grade 2, 12 mm (3-2-1), clear margins, no emboli, HER2 negative. 3 nodes examined, 0 positive (3N0/3). Ki-67 at 5%.<br>Left breast radiotherapy from xx/xx to xx/xx with boost.<br>Initiation of hormonal therapy (anastrozole) on xx/xx/xxxx (normal bone mineral density). | T1N0M0<br>Stage I |
| XX/XXX: Right breast carcinoma, cT4c (9 cm upper inner quadrant) N0 M1, invasive ductal carcinoma grade 2, ER 100%, PR 20%, HER2 negative, de novo metastatic to bone and stomach, with suspicious pulmonary nodules and a hepatic nodule.<br>Gastric biopsy: Carcinoma cells compatible with breast origin.<br>Participation in the PADA-1 study.<br>Start of letrozole + palbociclib on XX/XX/XXXX.<br>XX/XX: Bone progression, no visceral lesions, asymptomatic; fulvestrant proposed. | T4cN0M1<br>Stage IV |
| XXXX: Following a spontaneous miscarriage, the patient noticed a swelling in the right breast.<br>Bilateral mammography on XX/XX/XXXX:<br>Right breast: Increased density in the upper quadrants, extending over 3 cm. Axillary lymphadenopathy present.<br>Left breast: ACR 2.<br>Right breast lesion biopsy on XX/XX/XXXX: Invasive ductal carcinoma, grade I (2 + 2 + 1). ER 100%, PR 40%, HER2 score 0.<br>Staging work-up (chest-abdomen-pelvis CT, bone scan): No visceral or bone metastases detected.<br>Breast MRI on XX/XX/XXXX:<br>Right breast: Findings suggestive of multifocal carcinoma with inflammatory presentation. Right axilla involvement with at least 4 lymph nodes. Presence of a suspicious subcentimeter lymph node in the right internal mammary chain.<br>Left breast: ACR 1.<br>XX/XX/XXXX: 1st cycle EC100.<br>PET-CT on XX/XX/XXXX: Hypermetabolism in the primary lesion of the right breast, associated with right axillary and right internal mammary lymph node involvement. Uncertain left supraclavicular lymph node. No visceral or bone metastases.<br>XX/XX/XXXX: 2nd cycle EC100.<br>XX/XX/XXXX: 3rd cycle EC100.<br>PET-CT on XX/XX/XXXX: partial metabolic response.<br>XX/XX/XXXX – XX/XX/XXXX: 4th cycle EC100 + 4 cycles of Taxotere 75.<br>PET-CT on XX/XX/XXXX: Complete metabolic response.<br>XX/XX/XXXX: Right mastectomy with axillary lymph node dissection. | T4dN3bM0<br>Stage III |

|  |
| --- |
| Pathology report: Multiple residual foci (approximately 50) of invasive adenocarcinoma, 1–15 mm, located in the upper outer, lower outer, retroareolar and lower inner quadrants. Some tumour foci at the junction of the upper quadrants. Fibrous sequelae noted. No intraductal component. Clear surgical margins. No tumour infiltration of the dermis, subcutaneous tissue or nipple. 5 positive nodes out of 6 (1 with extracapsular extension). Stage ypT1cN2a. Sataloff grade: TcNd.<br>XX/XX/XXXX – XX/XX/XXXX: Radiotherapy, 50 Gy to right chest wall + axillary, supra– and infraclavicular lymph node areas + right internal mammary chain.<br>XX/XXXX: Initiation of tamoxifen. |

